# Impact of tozinameran (BNT162b2) mRNA vaccine on kidney transplant and chronic dialysis patients: 3-5 months followup

**DOI:** 10.1101/2021.06.12.21258813

**Authors:** Iddo Z. Ben-Dov, Yonatan Oster, Keren Tzukert, Talia Alster, Raneem Bader, Ruth Israeli, Haya Asayag, Michal Aharon, Ido Burstein, Hadas Pri-Chen, Ashraf Imam, Roy Abel, Irit Mor-Yosef Levi, Abed Khalaileh, Esther Oiknine-Djian, Aharon Bloch, Dana G. Wolf, Michal Dranitzki Elhalel

## Abstract

**Background:** Determining the humoral immunogenicity of tozinameran (BNT162b2) vaccine in patients requiring chronic renal replacement therapy, and its impact on COVID-19 morbidity several months after vaccination, will guide risk assessment and subsequent changes in vaccination policy.

**Methods:** In a prospective post-vaccination cohort study with up to 5 months follow-up we studied outpatient dialysis and kidney transplant patients and respective healthcare teams. Outcomes were anti S1/S2 antibody response to vaccine or infection and infection rate during followup.

**Results:** 175 dialysis patients (40% women, 65±15 years), 252 kidney transplant patients (33% women, 54±14 years) and 71 controls (65% women, 44±14 years) were followed. Three months or longer after vaccination we detected anti S1/S2 IgG antibodies in 80% of dialysis patients, 44% of transplant recipients and 100% of controls, whereas respective rates after infection were 94%, 75% and 100%. Predictors of non-response were older age, diabetes, history of cancer, lower lymphocyte count and lower vitamin-D levels. Factors associated with lower titers in dialysis patients were modality (hemodialysis vs peritoneal) and high serum ferritin levels. In transplant patients, hypertension and higher calcineurin or mTOR inhibitor drug levels were linked with diminished antibody response. Vaccination associated with fewer subsequent infections (HR=0.23, p<0.05). Moreover, higher antibody titers associated with fewer events, HR 0.41 for each unit increased in log_10_titer (p<0.05).

**Conclusions:** Dialysis patients, and more so kidney transplant recipients, mounted reduced antibody response to COVID-19 mRNA vaccination, and lesser humoral response associated with more infections. Measures to identify and protect non-responsive patients are urgently required.

**Significance:** Reports on the humoral immunogenicity of SARS-CoV-2 mRNA vaccines in patients with end stage renal disease are scarce, and association with subsequent COVID-19 morbidity is unknown. In this cohort study that included 175 patients treated with dialysis, 252 kidney transplant recipients and 71 control volunteers, the proportion achieving an antibody response was time- and group-dependent, reaching 80%, 44% and 100% at 3 months post prime inoculation. Personal history of vaccination, positive antibody responses and antibody titers associated with significantly lower risk of COVID-19 infection. Thus, in patients with end stage renal disease, SARS-CoV-2 antibody testing may be warranted after vaccination, to identify non-responders at higher risk for disease.

## Introduction

Vaccinations, both passive and active, changed the natural course of many infectious diseases, affecting both spread and severity. From smallpox through rabies, tetanus and measles, to hepatitis B, as well as many other viral and bacterial infections, vaccines have either eradicated diseases or eliminated the risk for pandemic or endemic catastrophes. Therefore, efforts to develop effective vaccination against COVID-19 started as soon as the impact of this coronavirus on global public health was appreciated. Impressively, effective vaccines were developed and approved at a record time, within less than a year. These active vaccines offer hope for controlling disease spread and reducing illness severity and death rates. Various vaccines are already in use, and data emerging form real-world practice (as from Israel^1^ and other countries) suggests effectivity in prevention of severe disease^2^.

The COVID-19 pandemic brought new challenges to the general population and even more so to patients with end-stage renal disease (ESRD), many of whom have comorbidities now acknowledged as risk factors for severe COVID-19. Aside from logistic challenges summarized elsewhere^3^, these patients, both dialysis-treated and kidney transplant patients, face morbidly and mortality risks that are significantly higher than the general population^4, 5^. Unfortunately, it is known from existing vaccine preparations such as anti-hepatitis B virus and the influenza^6-9^, that both dialysis and kidney transplant patients require higher doses and repeat inoculations (i.e. periodic boosts) in order to achieve durable protection^10, 11^. This diminished response to vaccination is thought to be secondary to dysfunction of both B and T lymphocytes in ESRD patients, as well as to the immunosuppressant medications taken by transplant recipients (occasionaly also after resuming dialysis). Dendritic cell dysfunction, described in patients with ESRD^12^ likely compromises vaccination success. As the information available about COVID-19 vaccine effectiveness in dialysis and transplanted patients is limited and short-termed^13, 14^, we sought to prospectively follow antibody development in these patients, to compare them with healthy controls and to identify risk factors for diminished humoral response and disease.

## Methods

### Setting

Vaccination with tozinameran, Pfizer and BioNTech’s BNT162b2 vaccine, commenced in Israel on December 2020-12-20, and immunocompromised patients were of the first priority groups. Prior to initiation of vaccination we launched the COVID-19 mRNA Vaccine Immunogenicity in patients with end stage Renal Disease (COVIReD) prospective cohort study designed to investigate the long-term kinetics and implications of antibody response to COVID-19 vaccine and infection in this vulnerable population. We are characterizing the humoral response to COVID-19 infection and vaccination as well as disease occurrence among patients treated with maintenance dialysis, kidney transplant recipients and control subjects at Hadassah Medical Center, a two-campus tertiary medical center in Jerusalem, Israel. Jerusalem was the area with the highest COVID-19 prevalence in Israel^15, 16^ (and during the pandemic more than 4,500 COVID-19 patients were admitted to our institution). We recruited control subjects from amongst medical, nursing and administrative healthcare staff at the dialysis unit and/or the transplantation clinic.

### Clinical methods

The study has been approved the the Haddasah Medical Organization’s Helsinki Committee and has been conducted in accordance with the declaration of Helsinki. Written informed concent was obtained from participants. Blood samples were taken from dialysis and transplant patients during routine visits, after obtaining informed consent, at several designated time-windows during the vaccination period: at baseline (namely, before administration of the first vaccine dose), 10-20 days after the first vaccine, 2-6 weeks after the second vaccine inoculation (typically scheduled 21 days after the first inoculation) and 3 months or longer after the first vaccination. Most hemodialysis patients and controls provided samples at all or most time-points while transplant and peritoneal dialysis patients provided samples sporadically during their scheduled outpatient visits.

### Laboratory methods

All serum samples were tested at Hadassah’s clinical virology laboratory, using kits supplied by the Israeli Ministry of Health. Anti-SARS-2 IgG antibodies were quantified using LIAISON SARS-CoV-2 S1/S2 IgG (DiaSorin) and ARCHITECT SARS-CoV-2 IgG II (Abbott) immunoassays, and the former kit results are reported here. In addition, before as well as after vaccination we tested nasal swabs for viral RNA using qPCR assays when clinically or epidemiologically indicated, but did not perform routine PCR screening except in staff. Inpatient and outpatient diagnoses, patient demographics and serology, virology and additional selected laboratory results (see **Table 1**) were extracted for the period between 2020-03-01 and up to patient vaccination, by the institution’s information systems.

**Table 1:**
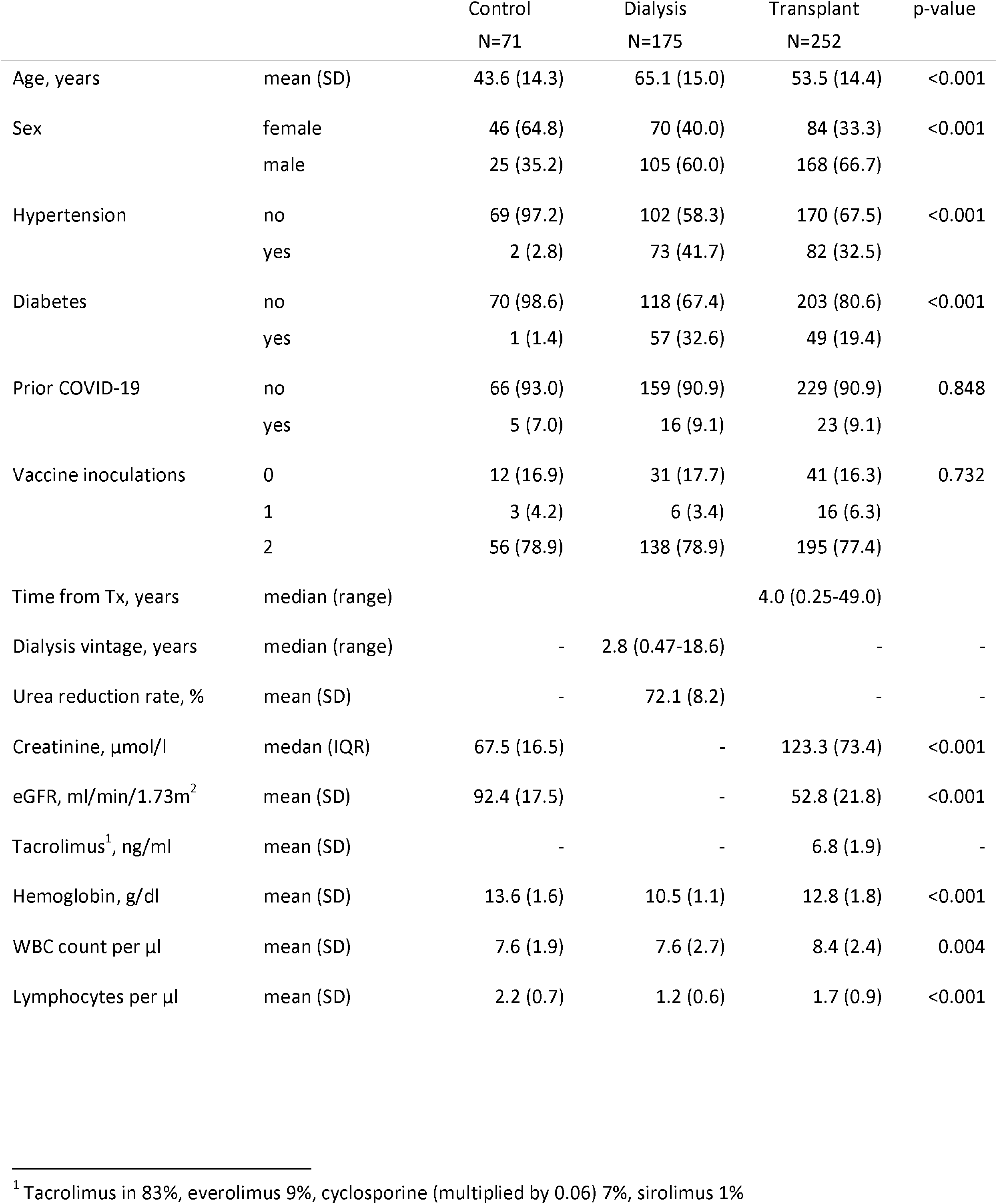
Demographic and clinical characteristics according to study group

### Statistical analysis

Deidentified clinical information and laboratory data and metadata were stored as spreadsheets, and processed using R base and related statistical packages. Clinical characteristics of study participants were summarized by group using means ± SD or medians and ranges, as appropriate. Right-skewed variables (e.g. antibody titers) were log_10_-transformed prior to statistical testing. Between-group differences in baseline clinical characteristics were assessed using ANOVA or chi squared. Statistical symbols embedded within plots reflect Mann-Whitney U test results. Potential clinical predictors of antibody response and disease (COVID-19 infection) were examined using mixed-effects linear (for antibody titers) and generalized linear (for dichotomous antibody results) and Cox proportional hazards models, respectively, using ‘lme4’^17^, ‘coxme’ (mixed effects cox models) and ‘survival’^18^ packages. Repeated measures were accounted for by including patient ID as a random effect in these models. Model outputs are presented using the ‘sjPlot’ package^19^. Predictions based on these models were generated and plotted using the ‘ggeffects’ package^20^. For Cox modeling of COVID-19 infection events with vaccination or serological test results as independent variables, the respective time-dependent covariates were constructed as suggested by Therneau et al using the ‘tmerge’ function of the ‘survival’ R/Bioconductor package^18^. We generated plots in R using the ‘ggplot2’^21^ and ‘NMF’^22^ packages.

## Results

One hundred and seventy-five patients treated with dialysis (152 hemodialysis and 23 peritoneal dialysis), 252 kidney transplant patients and 71 nephrology healthcase team control participants provided specimens for serological analyses. **Table 1** summarizes their demographic and clinical characteristics. Specimens were provided before, between and/or after administration of vaccine doses and COVID-19 infections, as desctibed in **Table S1** (online supplement). Infections occurred *prior* to vaccine availability in 7.0%, 9.1% and 9.1% of control, dialysis-treated and transplant participants, respectively. Twenty percent, 31% and 17% of these respective *prevalent* cases were identified solely via positive serology.

Of our study participants, 82-83% received at least 1 dose of tozinameran and 77-79% received 2 inoculations, while among participants who were not infected with COVID-19 prior to vaccine availability, complete vaccination rates were 83-85%. **Figure S1** (in the online supplement) shows the timeline of COVID-19 infections in our cohort, and of vaccination events, beginning at the epidemic onset.

### Serological results

**Figure 1a** and **Table S1** show the binary serological outcome among the 3 groups of participants at several occasions in relation to vaccination and disease. While 100% of control subjects generated borderline or above antibody levels when sampled 22-71 days after the first vaccination, only 80% of dialysis patients – and 44% of transplant recipients – achieved such levels when tested 3 months or more after the first dose (range 90-139 days). In unvaccinated COVID-19 survivors, the respective positive (incl. borderline) antibody rates were 100%, 94% and 75%. Predictors of lack of anti-spike IgG response after single or double vaccine inoculation were being a dialysis (odds ratio = 39, 95%CI 6.0-2.6×10^2^, p<0.001) or transplant patient (OR=7.1×10^2^, 95%CI 7.1-7.0×10^3^, p<0.00001) compared to control, shorter time interval between vaccination and testing (OR=0.97 per day, 95%CI 0.96-0.98, p<0.00001, **Figure 1b**), age above the median (58 years) (OR=3.7, 95%CI 1.6-8.8, p<0.01, and see **Figure 1c**), personal history of cancer (OR=5.9, 95%CI 1.3-27, p<0.05) or diabetes (OR=2.6, 95%CI 1.1-6.4, p<0.05), as well as lower lymphocyte count (OR=0.49, 95%CI 0.29-0.82, per 1,000 cells/µl, p<0.01) and (in subjects with available data) lower vitamin D levels (OR=0.94, 95%CI 0.89-1.00, per 1 ng/ml, p<0.05).

**Figure 1.**
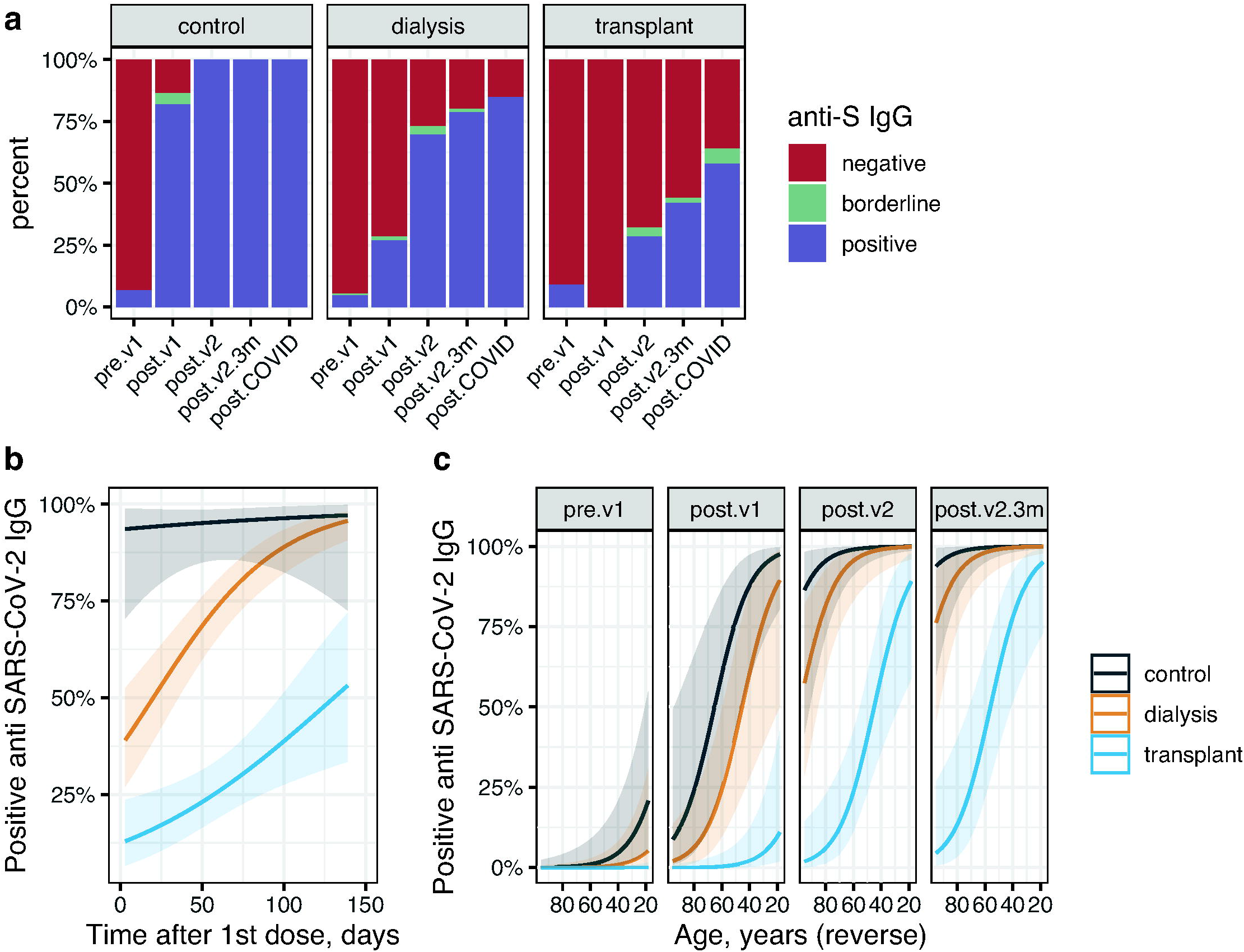
Qualitative results of anti S1/S2 SARS-CoV 2 IgG serology testing. **(a)** Distribution of categorical results among the study groups along the study time points. **(b)** Model prediction of IgG positivity rates in the 3 study groups according to the timing of testing in relation to the 1st vaccination. **(c)** Model prediction of IgG positivity rates in the 3 study groups at different time points according to participant’s age. Definitions: “pre.v1”, before vaccination; “post.v1”, between vaccinations; “post.v2”, up to 10 weeks after the 2nd vaccine; “post.v2.3m”, more than 10 weeks after the 2nd vaccine (3 months post 1st vaccine); “post.COVID”, after COVID-19 infection (regardless of vaccination status).

### Quantitative serological results

**Figure 2a** and **Table S2** show the numerical serological outcome among the 3 groups at several occasions, in relation to vaccination and disease. The quantitative titers parallel the qualitative results presented above. Linear mixed effects models indicated that in addition to the timepoint of testing, IgG titers depended on the study group, being 0.54 log_10_ lower values in dialysis patients and 1.29 log_10_ lower in transplant patients, compared to controls (both p<0.00001). Log_10_ titers were 0.01 lower per year of age (p<0.00001). In addition, higher lymphocyte counts (0.12 log_10_ per 1,000 cells/µl, p<0.01) and lower ferritin levels (0.03 log_10_ per 100 ng/ml, p<0.05) were linked with higher antibody titers. **Figure 2b** depicts antibody titers in participants that were vaccinated twice according to the time elapsed from the prime dose. Predictions based on linear mixed effects models (with time post vaccination introduced as an independent variable using splines and the time*group interaction also included in the model) are shown in **Figure 2c** and **2d**. A plateau is notable to emerge in controls and dialysis patients at ∼55 days post 1st vaccination, while in transplant recipients a mild incline may persist beyond this period.

**Figure 2:**
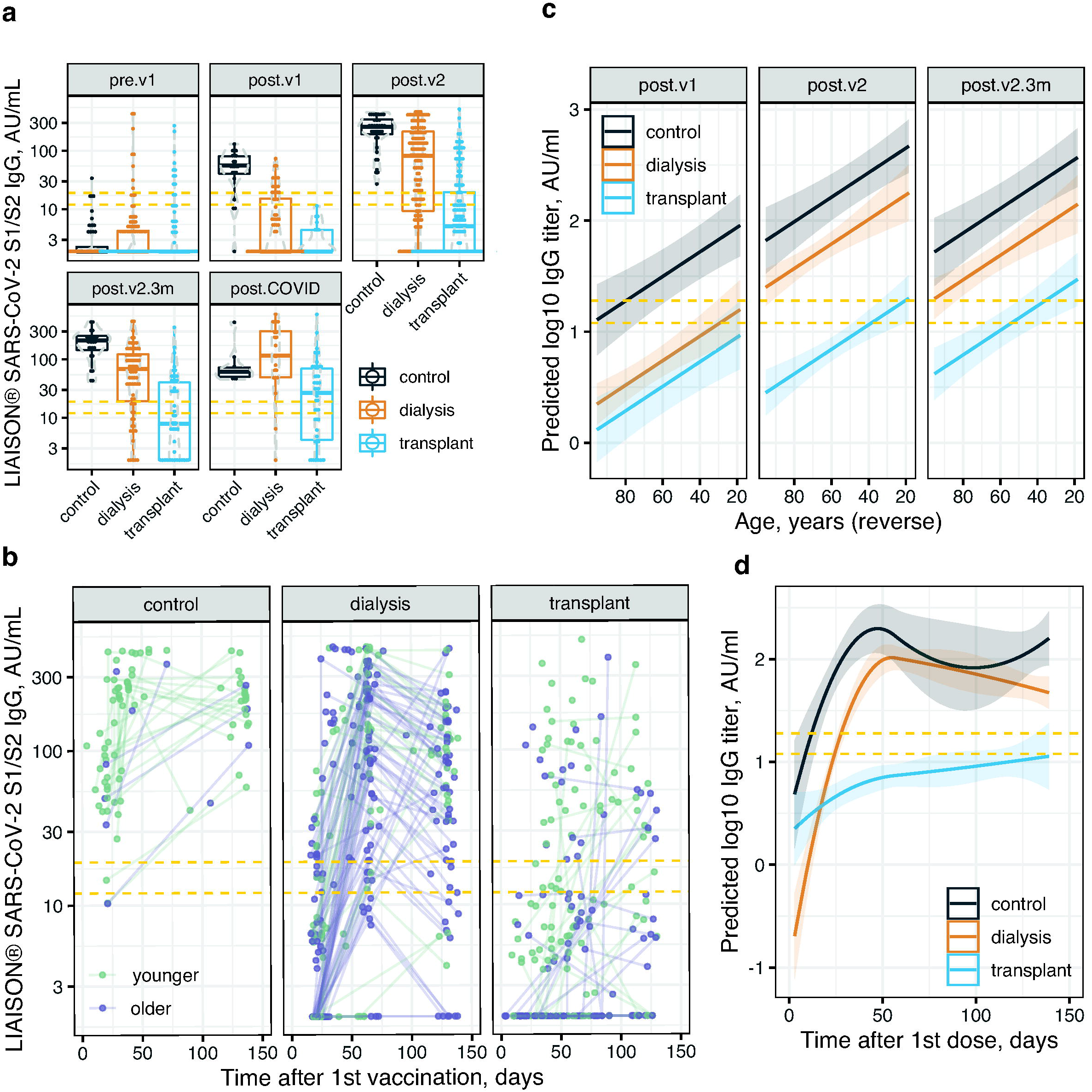
Numerical results of anti S1/S2 SARS-CoV 2 IgG serology testing. **(a)** Dot- and box-plots showing antibody titers (log scale) in the 3 study groups at 5 specified time points. **(b)** Scatter plot showing antibody titers (log scale) versus time after 1st vaccine in the 3 study groups. Repeat measurements from the same participant are connected with lines. **(c)** Model prediction of IgG titers versus age in the 3 study groups at different time points. **(d)** Model prediction of IgG titers versus time after 1st vaccination in the 3 study groups. See Figure 1 legend for time point definition. The dashed yellow lines represent the borderline titer range.

### Group-specific associations

In transplant recipients, the step-up in antibody titers after the 2nd vaccination was significantly greater in younger (<55 years) compared to older patients (**Figure S2**). In fact, 3 months or longer after the prime dose, 60% of younger patients but only 24% of older transplant patients had a categorically positive titer of antibodies. Time from transplantation was associated with antibody titers in a non-linear manner; compared to patients transplanted 6-18 months before antibody testing (having lowest levels), recipients less than 6 months after transplantation had 0.92 log_10_ higher titers (p<0.01) and recipients of 18 months or longer duration had 0.30 log_10_ higher titers (p<0.01). In models including age, timing of antibody testing and time after transplantation, titers were 0.20 log_10_ lower in patients with diagnosis of hypertension (p<0.05) and 0.51 log_10_ lower with diagnosis of anemia (p<0.05). The aggregated immunosuppressive drug level – tacrolimus, cyclosporin A (transformed by multiplying by 0.06), everolimus and sirolimus – was also linked with lower titers (0.063 log_10_ per ng/ml, p<0.05, **Figure S2c**), however this association was not significant when accounting for time after transplantation (0.055 log_10_ lower per ng/ml, p=0.08). Serum creatinine and estimated glomerular filtration rate (CKD-EPI equation) did not associate with titers.

In dialysis patients, unique predictors of antibody titers were: (1) modality; peritoneal dialysis patients had 0.41 higher log_10_titer compared to haemodialysis patients (adjusted for age and time post vaccination, p<0.01). (2) Ferritin levels were linked with lower log_10_titers (0.03 per 100 ng/ml, p<0.05) (Figure S3). Conversely, dialysis vintage, comorbid conditions and averaged urea reduction rate (in hemodialysis patients) were not associated with antibody titers.

### Occurrence of COVID-19

Ninety six study participants had COVID-19 infection before or after the vaccination period. Vaccine inoculations, introduced as time varying covariates, are seen in **Figure 3a** (and **Figure S4a**) to inversely associate with COVID-19 infection risk, after vaccine availability. This relationship was marginally stronger in controls compared to both ESRD groups (p-values for the interactions ∼0.1), but was significantly weaker in older compared to younger subjects (**Figure 3b** and **Figure S4b**), possibly owing to lower baseline risk in this subgroup (p-value for the interaction 0.013).

**Figure 3:**
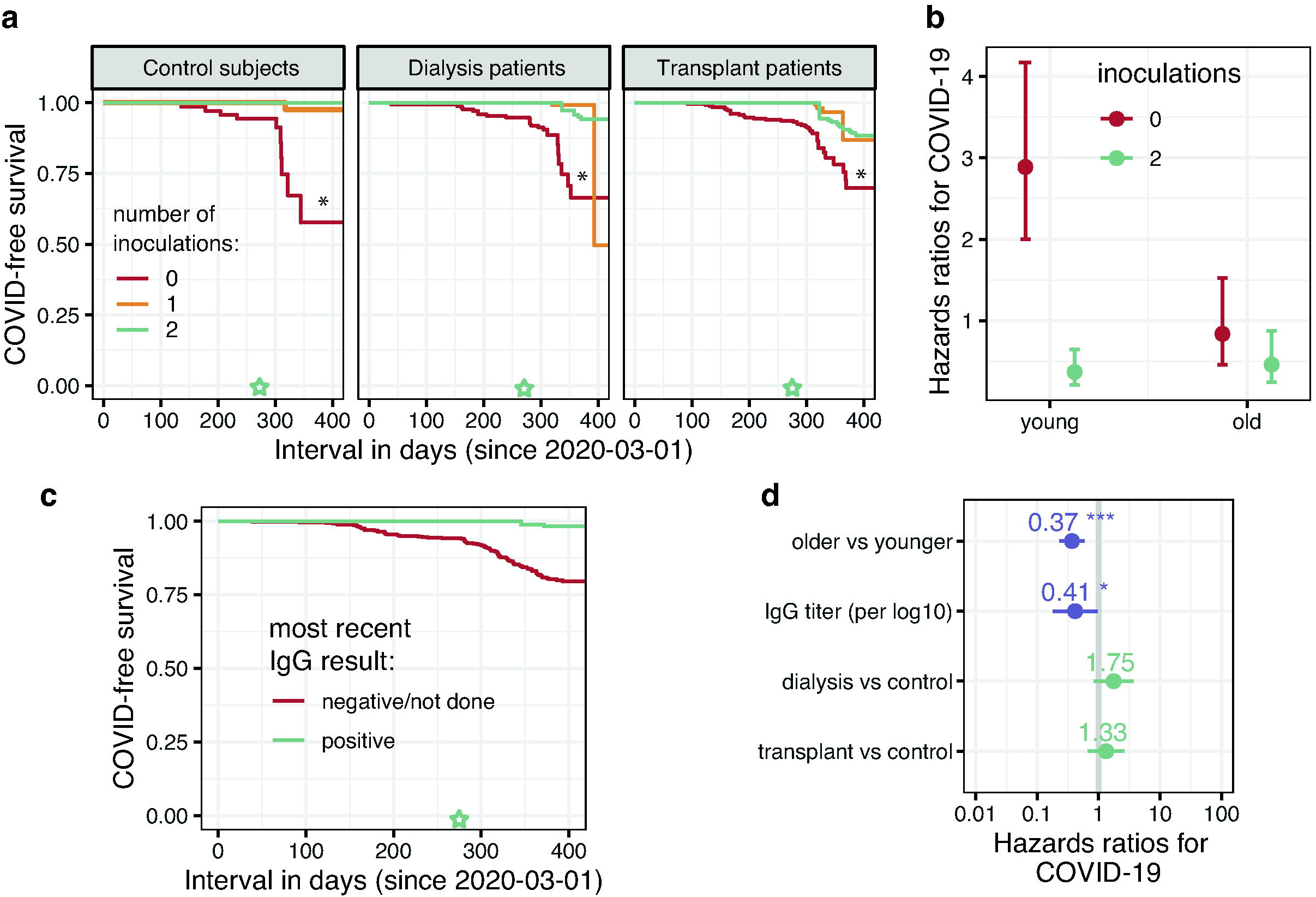
Associations between vaccine inoculation, anti S1/S2 IgG test results and COVID-19 infection. **(a)** Probability of COVID-19 infection from epidemic onset according to vaccination status as a time varying covariate (see Methods), split by study group. The inset shows the unspilt curves (all study groups). *, p<0.05 for 2 vs. 0-1 inoculations. **(b)** Risk of COVID-19 infection by inoculation status and age group. **(c)** COVID-19 events presented as Kaplan Meier curves according to IgG serology status. **(d)** Determinants of COVID-19 risk in a model including IgG titer as a time varying covariate. In **(c)** and **(d)** he green star marks the beginning of the vaccination period.

Moreover, positive (including borderline) anti-SARS-CoV-2 IgG serology after vaccination was linked with lower risk of COVID-19 infection (**Figure 3c** and **Figure S4c**), hazards ratios 0.23 (95% CI 0.05-0.99). Likewise, quantitative IgG titers were linked with significantly lower COVID-19 risk (**Figure 3d** and **Figure S4d**). See additional analyses including mortality as supplementary **Supplementary Text** and **Figure S5**.

### Post-COVID-19 serology

SARS-CoV-2 S1/S2 IgG antibody titers after COVID-19 infection were lower compared to vaccine-driven antibodies in controls, but not in ESRD patients. In fact, in transplant recipients the opposite was true (**Figure 4**).

**Figure 4:**
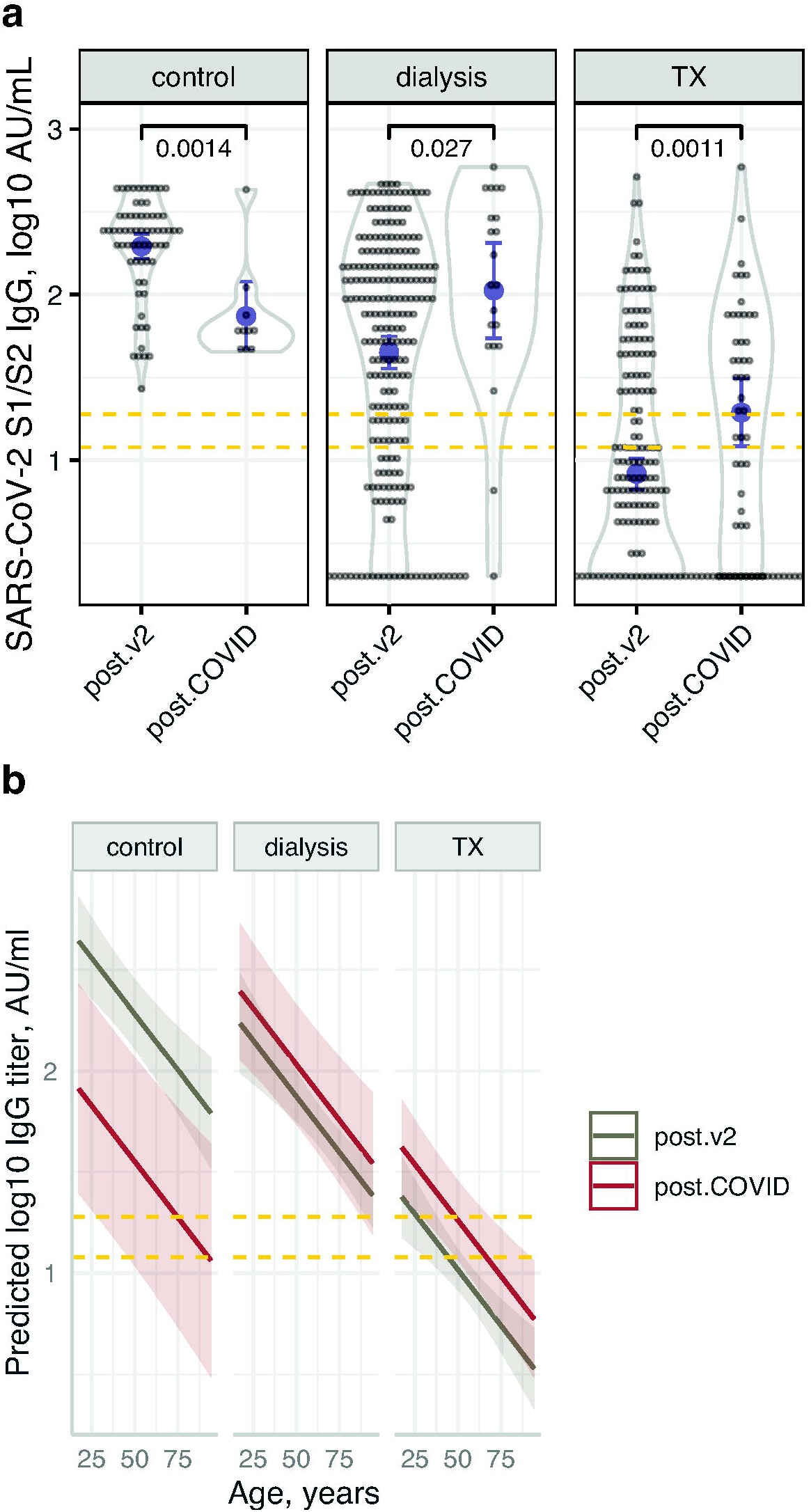
Comparison of post-vaccination with post-COVID-19 infection anti S1/S2 SARS-CoV 2 IgG titers. **(a)** Dot- and violin plots showing IgG titers post COVID-19 vaccination (“post.v2”) or infection (“post.COVID”). Also shown are the means and confidence limits (based on the t-distribution). **(b)** Model prediction of IgG titer versus age post-infection and post-vaccination in the 3 study groups, showing higher predicted titers post vaccine in controls but not in the ESRD groups.

## Discussion

This prospective study was designed to follow serum antibody levels after COVID-19 vaccination and/or disease in patients requiring chronic renal replacement therapy, and to estimate associations of vaccine inoculation and the resulting humoral response with subsequent COVID-19 infections. Healthcare team personnel (nursing, medical, assisting and administrative staff, working in the respective dialysis units and/or in the transplantation clinic) served as controls. Anti-spike antibody positivity rates and levels did not differ between groups at baseline. However, dialysis and more so kidney transplant patients had significantly lower positive response rates and titers both after the prime administration and after the 2nd, compared to the staff. These findings are in line with previous studies in kidney dialysis and transplant patients (reported predominanltly as “letters to the editor”). Not previously reported, we found that vaccination was linked with lower risk of COVID-19 infection in all groups; however, the effect was smaller in the ESRD patients.

Prior to vaccination, positive serology was found in a small percentage (<10%) of participants, indicating past infection with SARS-CoV-2 virus. About one third of these infections were not detected or suspected previously, despite strict screening measures in dialysis patients – questioning for possible contacts with verified COVID-19 patients, for any risk factor for exposure (as recent flight, attending crowded event or living in neighborhoods with high rates of infection), and for symptoms that can suggest active infection, as well as actively measuring body temperature and repeated SARS-CoV-2 nucleic acid quantification by nasal swabs after possible exposure.

There are significant age differences between the control group and the dialysis patient group, which is compatible with the average age of dialysis patients. As significant differences of antibody levels were found in the vaccine’s clinical study, where older individuals had lower antibody titer (also observed among our study partitipants), we included age in the statistical models, and thus report a significant age-adjusted relationship between study group and antibody titers.

Only 32% of transplant patients and 73% of dialysis patients had a positive (including borderline) antibody test after the second vaccination, improving by 3 months to 44% and 80% respectively, yet still having significantly lower antibody titers compared with controls. This raised the question whether additional vaccine boosts or higher doses are needed in this patient population, as was previously reported for hepatitis B vaccine^10^.

Strikingly, many fully vaccinated transplat patients were identified with no detectable antibodies, and some developed severe COVID-19 after infection. Similar findings, of reduced response to two-dose regimen of tozinameran, were reported previously in dialysis patients^24-27^, kidney transplant recipients^28-30^, as well other receivers of solid-organ alografts^31-33^, although follow-up periods were usually much shorter than in our study and there were typically no investigations of links with infection.

During the study period, infection rates in Israel including Jeusalem dropped drastically. However, we found that COVID-19 infection following vaccination was *independently* associated with lower post-vaccination antibody titers, suggesting benefit from personal history of vaccination on top of herd immunity. This association was reported previously with neutralizing antibody assays^34^ but not with antibodies detected by common ELISA. This association has clinical and epidemiological importance; if we can identify individuals at high risk of infection, we can apply individualized protective measures, while easing on the general population, or introduce additional vaccination boosts for that specific vulnerable group. The findings that lower lymphocyte counts and higher serum levels of immunosuppressive medications are associated with lesser antibody response also suggest the need for additional vaccination for immunosuppressed patients. Intrigingly, controls mounted lower anti S1/S2 titers after infection compared to vaccination, while the reverse was true in ESRD patients. This can be in part explained in transplant patients by the observation that during and shorly after infection, immunosuppressant medication doese were typically reduced, except corticosteroids which were often temporarily increased.

There are several limitations to our study. This is a single center study, and may not represent the larger ESRD community when considering future vaccination policy, although we believe that the study is large enough to facilitate further research. A longer follow-up period can enrich the data on antibody titers, although the dramatic rapid decline in COVID-19 prevalence in Israel means that hopefully little new data regarding protectivity will be available. Additionally, COVID-19 infection was defined as positive PCR, regardless of symptoms, but routine screening was not done, and therefore we might have missed several asymptomatic patients. Lastly, we did not report in this study ongoing investigation of cellular immunity or non-IgG antibodies which could add to our understanding of the immune response and protection after vaccination.

In **conclusion**, we show that ESRD patients exhibit impaired humoral response to two doses of tozinameran, a prototype of mRNA vaccination, manifested either by negative ELISA or lower titers than those of the healthy controls. Of note, a small rise in antibody levels and proportion of responding patiens is evident after three months, suggesting a different time scale of the immune response. Additionally, we show inverse association of IgG titers with the risk of contracting COVID-19 after vaccination. We suggest testing immune-compromised patients for COVID-19 IgG antibodies in order to identify high-risk patients, and to expand research regarding the need for boost doses.

## Data Availability

Data is not available

## Author Contributions

- Study conception and design – IZB-D, AK, MDE, DGW
- Data acquisition including patient recruitment – KT, TA, RB, RA, HA, MA, IB, HP-C, AI, RA, IM-YL, EOD, AB
- Data analysis –IZB-D
- Data interpretation –IZB-D, YO, MDE
- Drafting the manuscript –YO, IZB-D, MDE
- Revising the manuscript – KT, AK

## Acknowledgments

None

## Supplemental Table of Contents

Table S1: Qualitative serological test results in the 3 study groups at different time points

Table S2: Quantitative serological test results in the 3 study groups at different time points

Figure S1: Timeline of COVID-19 infection and vaccination events split by study groups

Figure S2: Transplant group-specific associations with numerical results of anti S1/S2 SARS-CoV 2 IgG serology testing

Figure S3: Dialysis group-specific associations with numerical results of anti S1/S2 SARS-CoV-2 IgG serology testing

Figure S4: Further associations between vaccine inoculation, anti S1/S2 IgG test results and COVID-19 infection

Suppl. Text: Mortality

Figure S5: Mortality in COVID-19 infection events among study participants

