## Supplemental figures and text for "Impact of tozinameran (BNT162b2) mRNA vaccine on kidney transplant and chronic dialysis patients: 3-5 months followup"

Content

Table S1:	Qualitative serological test results in the 3 study groups at different time points

Table S2:	Quantitative serological test results in the 3 study groups at different time points

Figure S1:	Timeline of COVID-19 infection and vaccination events split by study groups

Figure S2:	Transplant group-specific associations with numerical results of anti S1/S2 SARS-CoV 2 IgG serology testing

Figure S3:	Dialysis group-specific associations with numerical results of anti S1/S2 SARS-CoV 2 IgG serology testing

Figure S4:	Further associations between vaccine inoculation, anti S1/S2 IgG test results and COVID-19 infection

Supplementary Text:	Mortality

Figure S5:	Mortality in COVID-19 infection events among study participants

| Table S1: **Qualitative serological test results in the 3 study groups at different time points** | | | | | | | | | |
| --- | --- | --- | --- | --- | --- | --- | --- | --- | --- |
|  | **Number of measurements** | | | **Proportion with positive or borderline result** | | | **Proportion with positive result** | | |
|  | control | dialysis | transplant | control | dialysis | transplant | control | dialysis | transplant |
| Pre-vaccine | 60 | 152 | 112 | 7% | 5% | 9% | 7% | 5% | 9% |
| Post-prime | 22 | 67 | 22 | 86% | 28% | 0% | 82% | 27% | 0% |
| Post-2nd | 40 | 148 | 137 | 100% | 73% | 32% | 100% | 70% | 28% |
| 3m.post-prime* | 26 | 80 | 50 | 100% | 80% | 44% | 100% | 79% | 42% |
| Post-COVID | 9 | 18 | 32 | 100% | 94% | 75% | 100% | 94% | 69% |
| Post-COV+v* | 1 | 8 | 17 | 100% | 75% | 53% | 100% | 75% | 47% |
| Post-bam* | 0 | 3 | 4 | — | 100% | 100% | — | 100% | 100% |
| *, “3m.post-prime” relates to 10 weeks after the 2nd vaccine (3 months after the 1st dose); “Post-COV+v” relates to testing after both COVID-19 infection and vaccination; “Post-bam” relates to testing after bamlanivimab treatment with or without vaccination. | | | | | | | | | |

| Table S2: **Quantitative serological test results in the 3 study groups at different time points** | | | | | | | | | |
| --- | --- | --- | --- | --- | --- | --- | --- | --- | --- |
|  | **Number of measurements** | | | **Means of IgG titers** | | | **Medians of IgG titers** | | |
|  | control | dialysis | transplant | control | dialysis | transplant | control | dialysis | transplant |
| Pre-vaccine | 60 | 152 | 112 | 4 | 7 | 7 | 2 | 2 | 2 |
| Post-prime | 22 | 67 | 22 | 58 | 11 | 3 | 56 | 2 | 2 |
| Post-2nd | 40 | 148 | 137 | 256 | 130 | 29 | 256 | 82 | 5 |
| 3m.post-prime* | 26 | 80 | 50 | 204 | 89 | 30 | 210 | 68 | 8 |
| Post-COVID | 9 | 18 | 32 | 63 | 186 | 46 | 58 | 117 | 33 |
| Post-COV+v* | 1 | 8 | 17 | 431 | 227 | 84 | 431 | 161 | 10 |
| Post-bam* | 0 | 3 | 4 | — | 334 | 133 | — | 317 | 106 |
| Titers are presented as IU/mL as reported from the LIAISON SARS-CoV-2 S1/S2 IgG (DiaSorin) test results.  *, “3m.post-prime” relates to 10 weeks after the 2nd vaccine (3 months after the 1st dose); “Post-COV+v” relates to testing after both COVID-19 infection and vaccination; “Post-bam” relates to testing after bamlanivimab treatment with or without vaccination. | | | | | | | | | |

Figure S1: **Timeline of COVID-19 infection and vaccination events split by study groups**

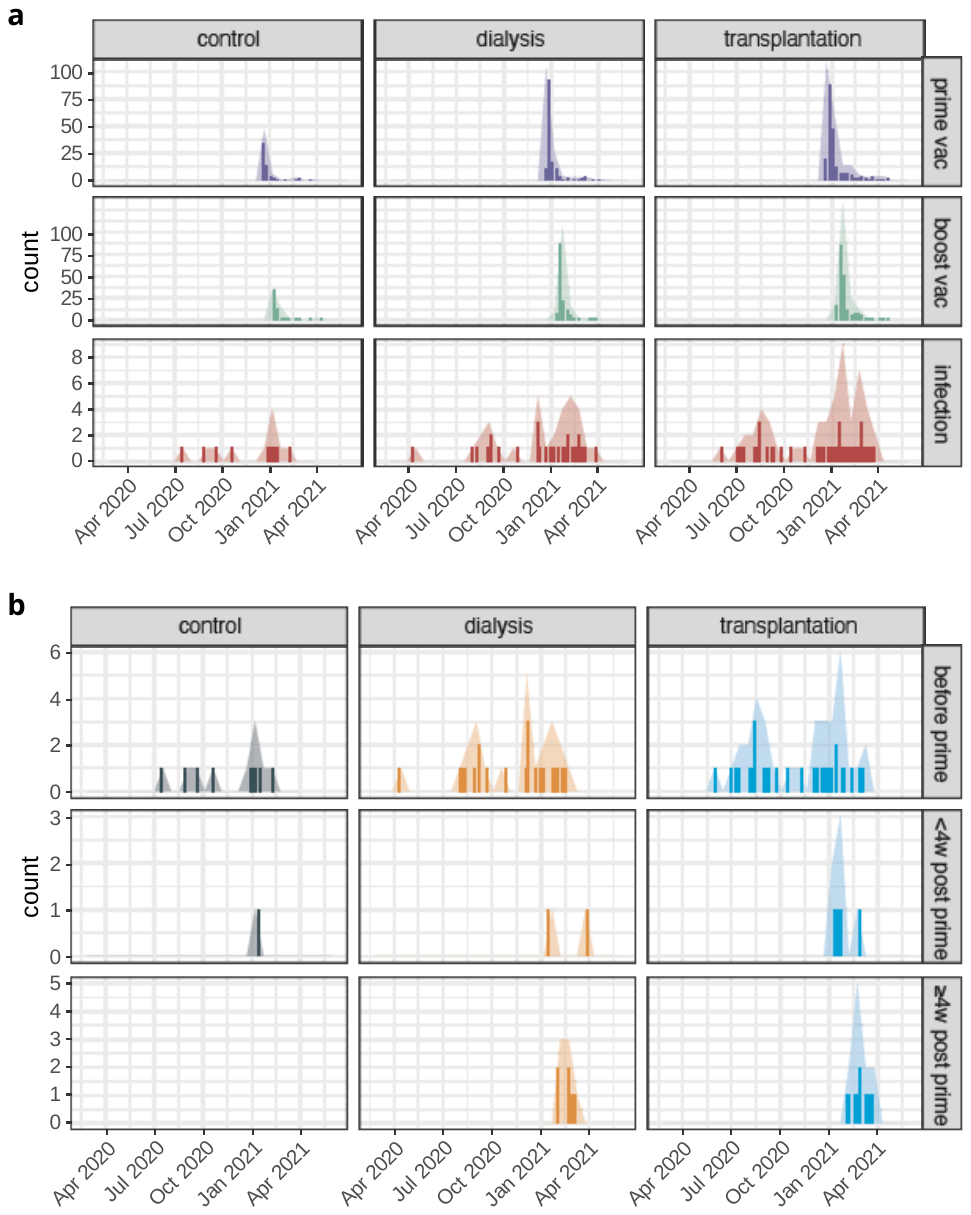

(**a**) Infections are shown in the lower panel, prime vaccination in the upper panel and 2nd vaccination in the middle panel. (**b**) Timeline of COVID-19 infections split by study groups. The upper panel shows events that occurred before vaccination; the middle panel depicts events within 4 weeks of 1st vaccination and the bottom panel shows infections occurring 4 weeks or longer after the prime inoculation.

Figure S2: **Transplant group-specific associations with numerical results of anti S1/S2 SARS-CoV 2 IgG serology testing**

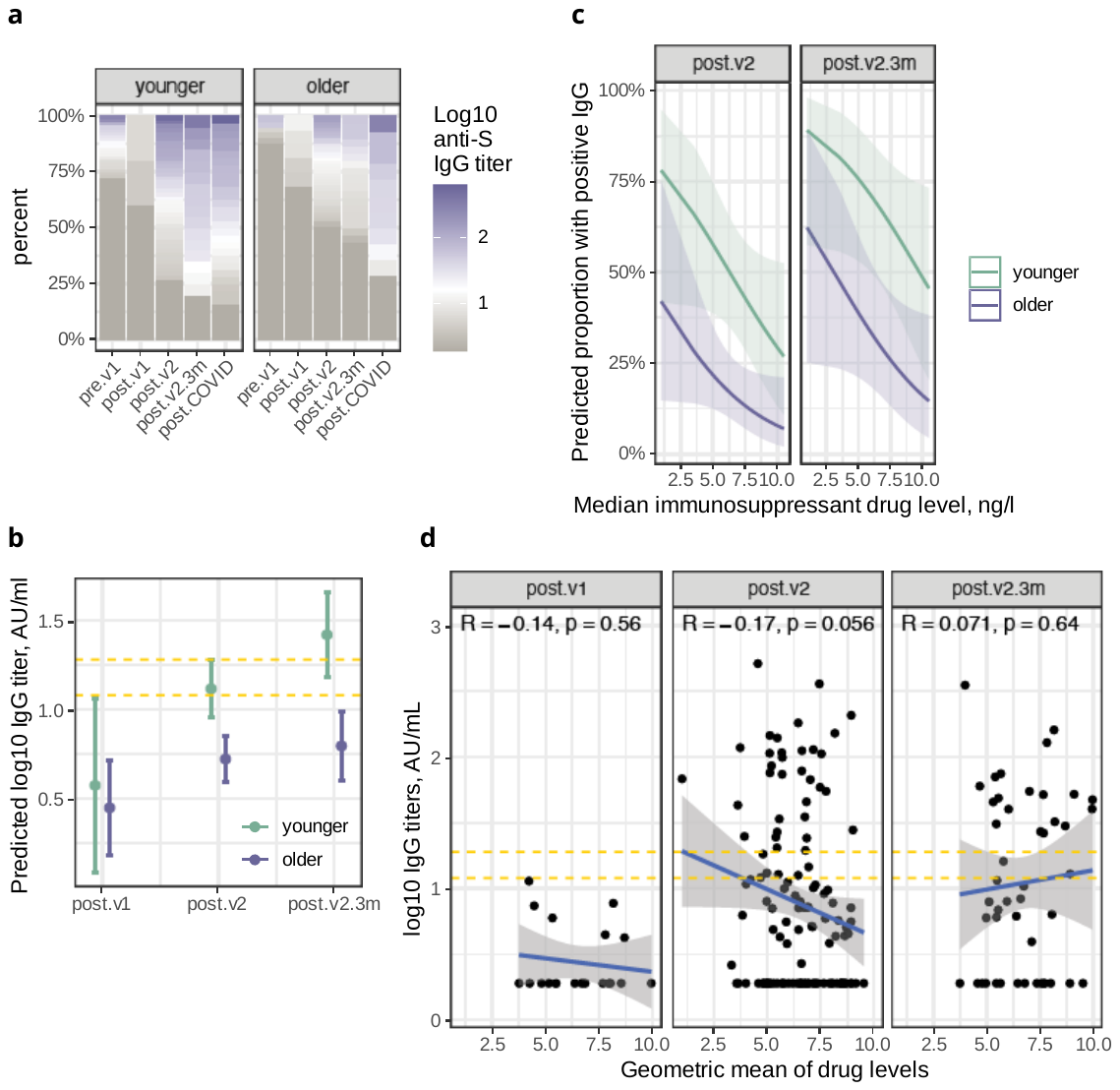

(**a**) Distribution of the titer results (log10 scale) in younger (<55 years) and older transplant patients across the study time points. Gray shades represent negative titers (<12 AU/mL), white represents borderline values and blue-violet shades represent positive titers (>19 AU/mL). (**b**) Model prediction of log10 antibody titers vs the immunosuppressant drug level by age group and study time point. Tacrolimus, cyclosporine (multiplied by 0.06), everolimus and sirolimus were aggregated for this analysis. (**c**) Model prediction of log10 antibody titers according to the interaction between study time point and age group. (**d**) Scatter plot showing antibody titers (log10) versus the immunosuppressant drug level. Dashed yellow lines represent the borderline titer range.

Figure S3: **Dialysis group-specific associations with numerical results of anti S1/S2 SARS-CoV 2 IgG serology testing**

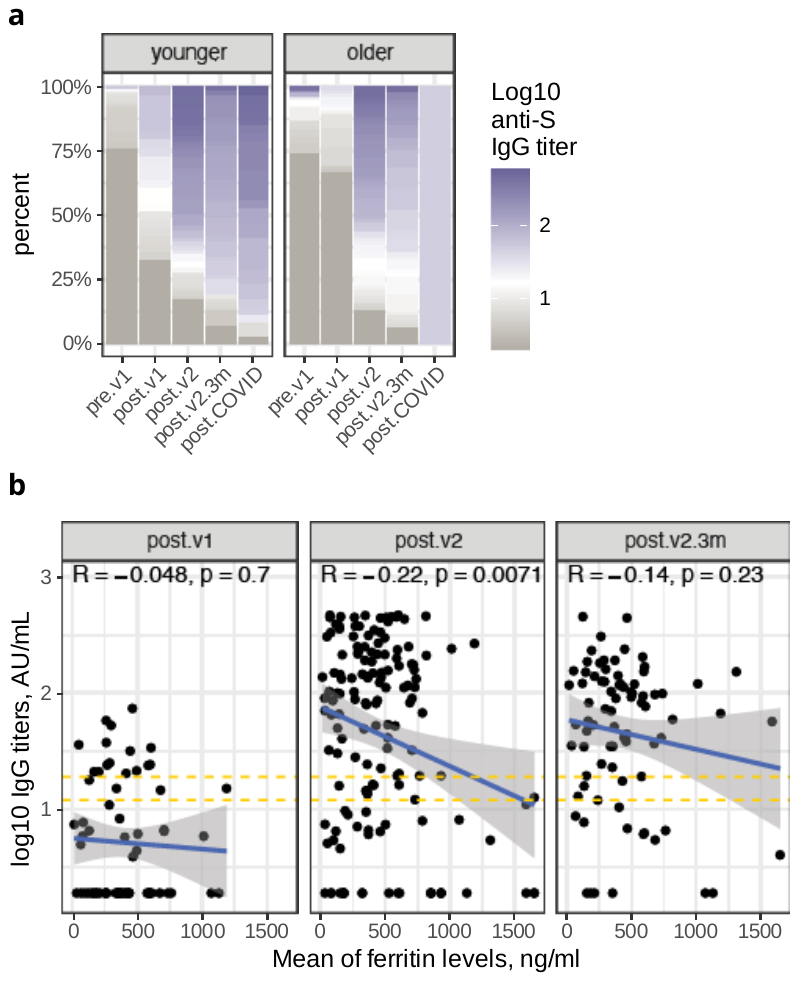

(**a**) Distribution of the titer results (log10 scale) in younger (<65 years) and older dialysis patients across the study time points. Gray shades represent negative titers (<12 AU/mL), white represents borderline values and blue-violet shades represent positive titers (>19 AU/mL). (**b**) Scatter plot showing antibody titers (log_10_) versus the mean serum ferritin level. Dashed yellow lines represent the borderline titer range.

Figure S4: **Further** **associations between vaccine inoculation, anti S1/S2 IgG test results and COVID-19 infection.**

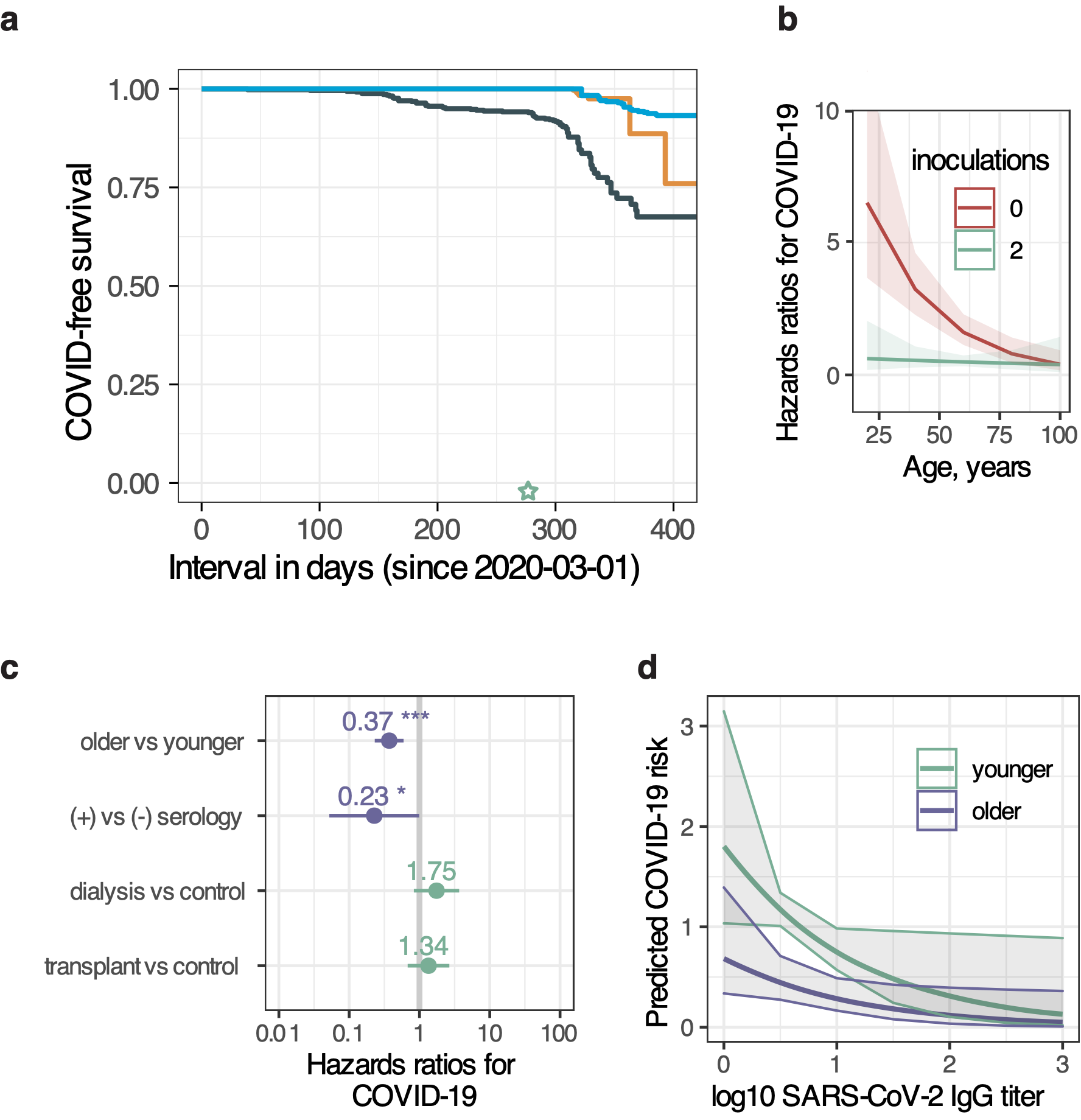

(**a**) Probability of COVID-19 infection from epidemic onset according to vaccination status as a time varying covariate (see Methods), **not** split by study group. (**b**) Risk of COVID-19 infection by inoculation status and age. (**c**) Determinants of COVID-19 risk in a model including qualitative IgG test result as a time varying covariate. (**d**) Model predicted infection risk by antibody titer, in 2 age groups.

**Supplementary Text: Mortality**

After the vaccine became available, 15 participants have died (12 HD, 1 PD and 2 Tx patients). Of these, 5 (33%) died of COVID-19 (3 HD and 2 Tx patients^[1]^), and another HD patient died 2 months after COVID-19. Three of the above patients had received two doses of the vaccine prior to infection with COVID-19, and one patient received a single dose. In an analysis limited to events after vaccine availability (2021-12-20), COVID-19-free and overall survival were inversely associated with age (**Figure S4**, left). Both outcomes were closely associated with the study group (**Figure S4**, right). SARS-CoV-2 IgG antibody titer, as a time-varying covariate, was not associated with lower mortality.

Figure S5: **Mortality in COVID-19 infection events among study participants**

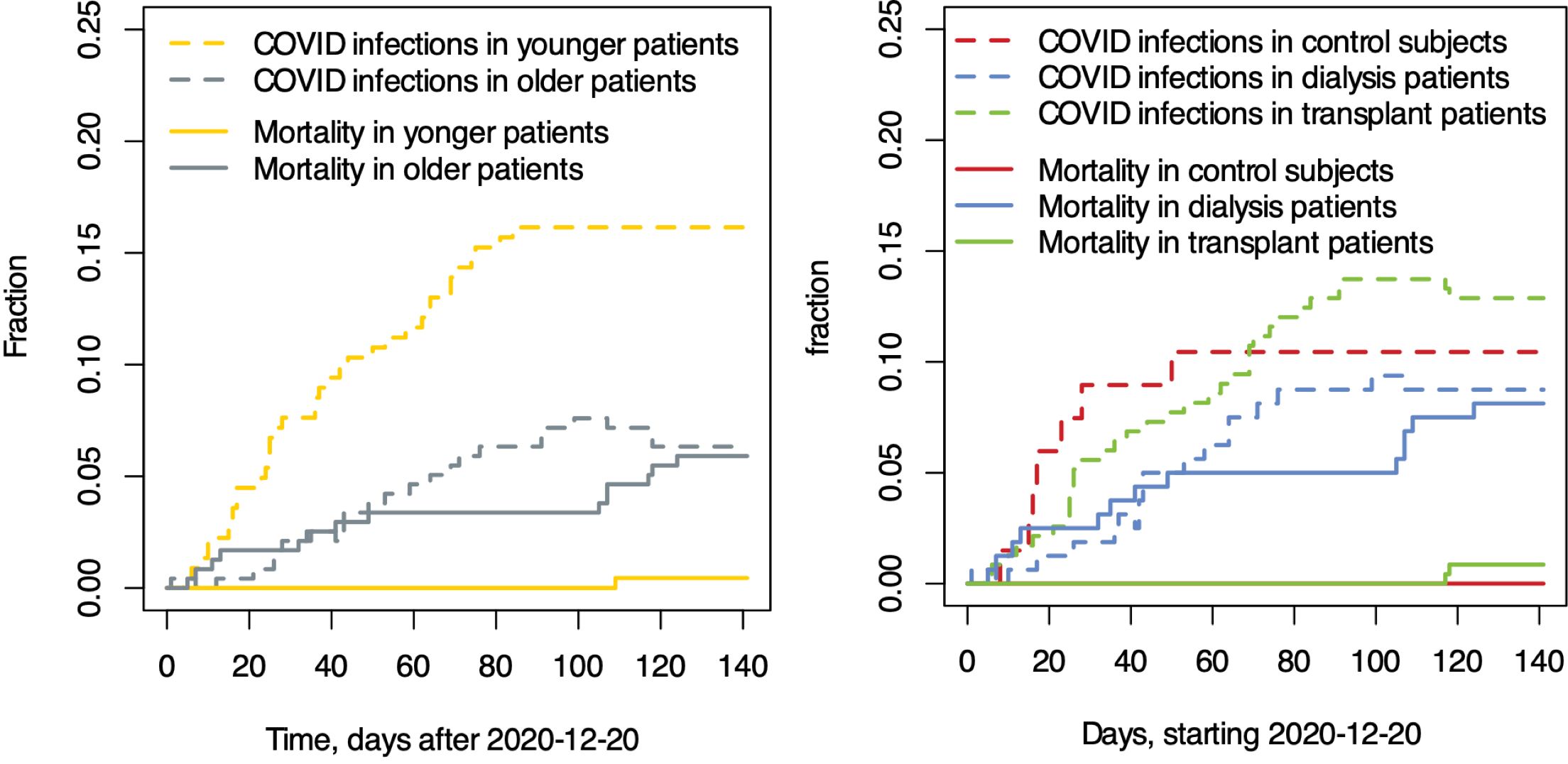

Kaplan Meier curves depicting mortality (all-cause) and COVID-19 infection events among study participants split by age group (**left**) or study group (**right**).

1. One patient, a female in her 60’s, had an antibody titer of 20.4 IU/mL (positive, ≥19 IU/mL) 64 days after prime vaccination which dropped to <3.8 IU/mL during disease (84 days post first vaccine inoculation) and died despite treatment with bamlanivimab at presentation. The second patient, a male in his 60’s, had an antibody titer of 7.2 IU/ml 74 days after the first vaccination, which rose up to 37.9 IU/mL shortly after presentation with COVID-19.
